# Travel time to health facilities as a marker of geographical accessibility across heterogeneous land coverage in Peru

**DOI:** 10.1101/19007856

**Authors:** Gabriel Carrasco-Escobar, Edgar Manrique, Kelly Tello-Lizarraga, J. Jaime Miranda

## Abstract

The geographical accessibility to health facilities is conditioned by the topography and environmental conditions overlapped with different transport facilities between rural and urban areas. To better estimate the travel time to the most proximate health facility infrastructure and determine the differences across heterogeneous land coverage types, this study explored the use of a novel cloud-based geospatial modeling approach and use as a case study the unique geographical and ecological diversity in the Peruvian territory. Geospatial data of 145,134 cities and villages and 8,067 health facilities in Peru were gathered with land coverage types, roads infrastructure, navigable river networks, and digital elevation data to produce high-resolution (30 m) estimates of travel time to the most proximate health facility across the country. This study estimated important variations in travel time between urban and rural settings across the 16 major land coverage types in Peru, that in turn, overlaps with socio-economic profiles of the villages. The median travel time to primary, secondary, and tertiary healthcare facilities was 1.9, 2.3, and 2.2 folds higher in rural than urban settings, respectively. Also, higher travel time values were observed in areas with a high proportion of the population with unsatisfied basic needs. In so doing, this study provides a new methodology to estimate travel time to health facilities as a tool to enhance the understanding and characterization of the profiles of accessibility to health facilities in low- and middle-income countries (LMIC), calling for a service delivery redesign to maximize high quality of care.

## 1. INTRODUCTION

Despite growing consensus to combat inequalities in the accessibility to healthcare around the world, large disparities in healthcare accessibility remain as a problem in countries with an ongoing rural-to-urban transition. According to the ‘Tracking Universal Health Coverage: 2017 Global Monitoring Report’, half of the worldwide population lacks essential health services (World Health Organization & World Bank, 2017). To overcome the disadvantage of marginalized populations, the international community through the United Nations (UN) have stated 17 Sustainable Development Goals (SDG) targeted by 2030 (UN General Assembly, 2015). From these goals, the interface between goal 3, — ‘Good health and well-being’,; and goal 10, — ‘Reduced inequalities’, play an important role to foster and couple endeavors towards ensured access to healthcare services.

The broad term ‘accessibility’, when referring to healthcare, focuses on multiple domains such as the provision of healthcare facilities, supply chain, quality and effective services, human resources, and on the demand side, health-seeking behaviors (Agbenyo, Marshall Nunbogu, & Dongzagla, 2017; Peters et al., 2008). All these characteristics pointed to the ability of a population to receive appropriate, affordable and quality medical care when needed (Kanuganti, Sarkar, Singh, & Arkatkar, 2015). Importantly, in rural and high poverty areas the most common reasons that prevents the access to healthcare are the geographical accessibility, availability of the right type of care, financial accessibility, and acceptability of service (Al-Taiar, Clark, Longenecker, & Whitty, 2010; Peters et al., 2008). This study focuses on the travel time to health facilities as an important component of the geographical (or physical) accessibility to healthcare.

Several studies in developing countries report that geographical accessibility is the main factor that prevents the use of primary healthcare (Al-Taiar et al., 2010; Kanuganti et al., 2015; Müller, Smith, Mellor, Rare, & Genton, 1998; Noor, Zurovac, Hay, Ochola, & Snow, 2003; Perry & Gesler, 2000; Stock, 1983), and not only conditions the ability of the population for health seeking, but also the capacity of the health system to implement prevention and control strategies with adequate coverage. However, fewer studies have explored the heterogeneity in geographical accessibility across areas with contrasting land coverage (Bashshur, Shannon, & Metzner, 1971; Comber, Brunsdon, & Radburn, 2011), i.e. the marked variation in the topography and environment conditions overlapped with different transport facilities between rural and urban areas that may influence the geographical accessibility across these areas. The geographical accessibility to health services has a direct impact on health outcomes since determine the timely response to patients that seek care, community-based campaigns (i.e. vaccination, iron supplements to combat anemia, etc.), or deliver first response to accidents or natural disasters.

Previous studies highlighted the importance of geographical or physical accessibility using a variety of methods (Comber et al., 2011; Delamater, Messina, Shortridge, & Grady, 2012; Huerta Munoz & Källestål, 2012; Ouko, Gachari, Sichangi, & Alegana, s. f.). The emergence of ‘Precision Public Health’ driven by estimates of a wide range of health indicators at a high spatial resolution is defined as the use of the best available data to target more effectively and efficiently interventions of all kinds to those most in need (Chowkwanyun, Bayer, & Galea, 2018; Dowell, Blazes, & Desmond-Hellmann, 2016; Horton, 2018; Tatem, 2018). This approach may be favorable since traditionally government’s reports aggregates data at administrative units, in a way that obscure the prioritization of resources. A recent study used a precision public health approach to estimate the geographical accessibility to major cities (Weiss et al., 2018), however, this approach has not yet been used for estimating the geographical accessibility to health facilities in developing countries.

This study sought to estimate the travel time to the most proximate health facility in rural and urban areas across heterogeneous land coverage types in Peru as a means to help resources prioritization, disease surveillance, as well as prevention and control strategies. Multiple sources of geospatial data were fitted with a novel cloud-based geospatial modeling approach (Weiss et al., 2018) to produce high-resolution (30 m) estimates of travel time to the most proximate health facility across the country. These estimates were then compared between urban and rural settings and across 16 major land coverage types in Peru.

## 2. MATERIALS AND METHODS

### 2.1. Study design

Ecological study using the Peruvian registry of villages and health facilities to model the travel time required for individuals in each village to reach the most proximate health facility (shortest travel time) in a two-step process. First, a friction surface was computed. Several geospatial datasets (land coverage types, boundaries of restricted areas, roads infrastructure, navigable river networks, and topography) were used to construct a surface (i.e. raster or grid) of a given spatial resolution (i.e. 30m per pixel) where the value of each pixel (or cell) contains the time required to travel one meter in that given area. Secondly, this friction surface and the geolocation of the health facilities were used to infer the travel time to the the most proximate (lowest time) health facility using a cumulative cost function. As a result, the travel time estimate for the most proximate health facility was computed for the entire country. The computed values were then summarized in a 500m-radius from the geolocation of cities and villages; per district, province or department; by urban/rural areas; and across 16 major land coverage types defined by the Ministry of Environment (MEnv).

### 2.2. Study area

This study was conducted using nationwide data from Peru, located on the Pacific coast of South America. Peru encompasses an area of 1,285,216 Km^2^ and 32,162,184 inhabitants divided in 25 departments and 1,722 districts. Major ecological areas in the country are divided into the coast, highlands, and jungle (**Figure 1A**), however this study explore a higher granularity of ecological areas with more than 60 unique land coverage areas (**Supplementary Information 1**) that were officially classified in Peru. This classification was based on ecological, topographic, and climate characteristics, that in turn are important for the calculation of travel time since each category requires a different displacement effort.

**Figure 1.**
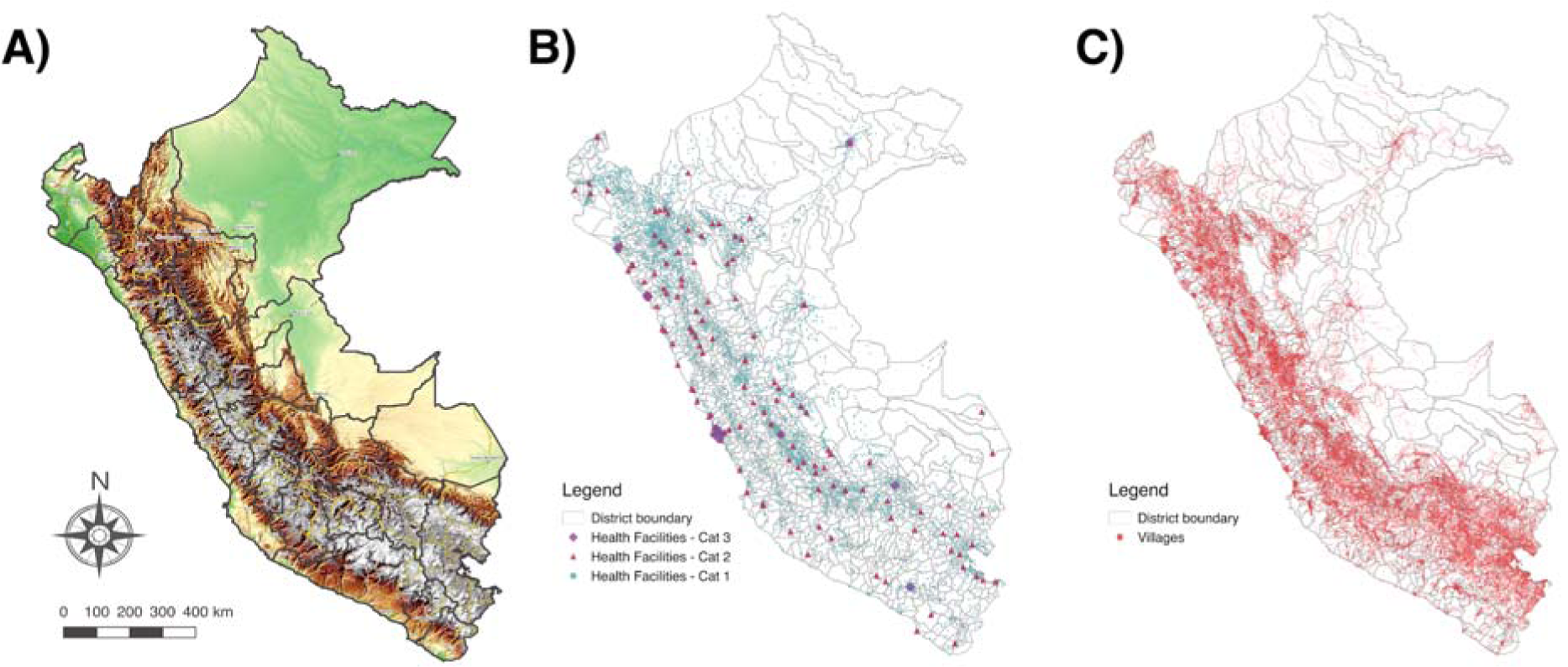
Study area. A) Major ecological areas (coast, andes, and jungle) in Peru. Solid lines represent the 25 Departments (administrative level 1). B) Spatial location of primary, secondary, and tertiary health facilities. C) Spatial location of villages. Maps were produced using QGIS, and the base map was obtained OpenTopoMap (http://www.opentopomap.org), under CC BY-SA 3.0.

### 2.3. Data Sources

The datasets were divided according to its use in the construction of the friction surface and the travel time map.

#### a) Friction surface construction

The land coverage types were derived from satellite images from MODIS MCD12Q1 product (Friedl et al., 2010). The MODIS collection includes seventeen land coverage types including urban and rural areas inferred by the spectral signature of the satellite images. The boundaries of the national protected natural areas were included using data provided by the MEnv. The road infrastructure in all districts was provided by the Peruvian Ministry of Transportation (MTrans), and the navigable river network was derived from the HydroSHEDS Flow Accumulation dataset (Lehner, Verdin, & Jarvis, 2006). The estimates of the friction surface (minutes required to travel one meter) were adjusted by the slope of the terrain. This means that, the travel time required to cross an area will be proportionally dependent to the slope of the terrain. The slope for each area was calculated using the SRTM Digital Elevation Data (Jarvis, Reuter, Nelson, & Guevara, 2008) produced by NASA.

#### b) Travel time estimation

The target locations used for the cumulative cost function were the health facilities (clinics) of the Ministry of Health (MH). This data was obtained from the geo-localization registry of health facilities (RENAES in spanish) (**Figure 1B**). The MH organize the health facilities in three categories according to the complexity of services they provide (from primary healthcare to specialized hospitals). The primary level includes basic health facilities with no laboratory, the secondary level includes health facilities with laboratory, and tertiary level includes hospitals and higher complexity services. Finally, travel time estimates were extracted for each city and village (**Figure 1C**). The most updated geo-localization of villages was provided by the Ministry of Education (MEd) in a recent census of cities and villages, and education facilities.

### 2.4. Data Analysis

#### a) Friction surface construction

The estimation of travel time were conducted in Google Earth Engine (GEE) (Gorelick et al., 2017). A surface grid was constructed using the information about land coverage, road infrastructure, and river network. All datasets were converted into aligned grids with a 30-meter resolution. Each dataset contained the information of the speed of movement in each feature. All the layers were then combined with the fastest mode of movement taking precedence (Km h^-1^). The speed assigned for each category of land cover were obtained from elsewhere (Weiss et al., 2018). A data transformation was conducted, so each pixel within the 2D grid contained the cost (time) to moving through the area encompassed in the pixel, herein referred to as ‘friction surface’. Slope adjustment was carried out using the Tobler’s Hiking Function (Tobler, 1993) and the speed was penalized (reduced) in urban and national protected areas to account for vehicular traffic and restricted displacement, respectively.

#### b) Travel time estimation

To calculate the travel time from the villages to the most proximate health facility, the cumulative cost function was used in GEE to generate the accessibility map. The cumulative cost function is a least-cost-path algorithm, briefly, all possible paths were analyzed iteratively and the weighted cost (in this case, weighted by time) was then minimized. The minimum travel time to the most proximate health facility was computed for each pixel in the grid, then the median travel time was summarized in a 500m-radius from the geolocation of each city or village (**Supplementary Information 2**). Values between the 5% and 95% percentile range were considered to avoid extreme values. Since a health facility could be located in the 30m^2^ corresponding to the pixel spatial resolution of the estimates, a baseline 10-minutes travel time was considered. The analysis was carried out for each health facility category. After GEE processing, all data outputs were imported and analyzed using R v.3.6.0 (R Development Core Team, R Foundation for Statistical Computing, Vienna, Australia).

The computed travel time was then summarized per district, province or department; by urban/rural areas; and across 16 major land coverage types defined by the MEnv. Urban/rural status was defined based on the MODIS land coverage satellite images (described previously in 2.3 Data Sources). To better detail the large diversity of land coverage types in Peru, a shortlist of 16 eco-regions provided by the MEnv (**Supplementary Information 1**) was used to summarize the travel time in these areas. In addition, the distribution of travel time relative to the proportion of population with unsatisfied basic needs (UBN) - a multidimensional poverty measurement developed by the United Nation’s Economic Commission for Latin America and the Caribbean (ECLAC) - per department was computed with data provided by the Ministry of Economy (MEco).

## 3. RESULTS

### Travel time to health facilities

For this study, we gathered geo-referenced data on 145,134 villages (**Figure 1B**) and 8,067 health facilities (**Figure 1C**) across the 1,722 districts in the Peruvian territory. The health facility density (number of health facilities divided by the total population) in Peru was 2.58 per 10,000 inhabitants with variations between major ecological areas, from 1.35 in the coast, 4.56 in the highlands, to 5.21 in the jungle.

Friction and travel time maps were reconstructed in Google Earth Engine using the described local datasets at a spatial resolution of 30 meters per pixel (**Supplementary Information 2**). Country-wide median travel time from each village to the most proximate health facility varies according to category: primary healthcare = 39 minutes (IQR=20 – 93), secondary healthcare = 152 minutes (IQR=75 – 251), and tertiary healthcare = 448 minutes (IQR=302 – 631).

Importantly, maximum travel time reached 7,819, 12,429, and 35,753 minutes for primary, secondary, and tertiary levels, respectively (**Figure 2**).

**Figure 2.**
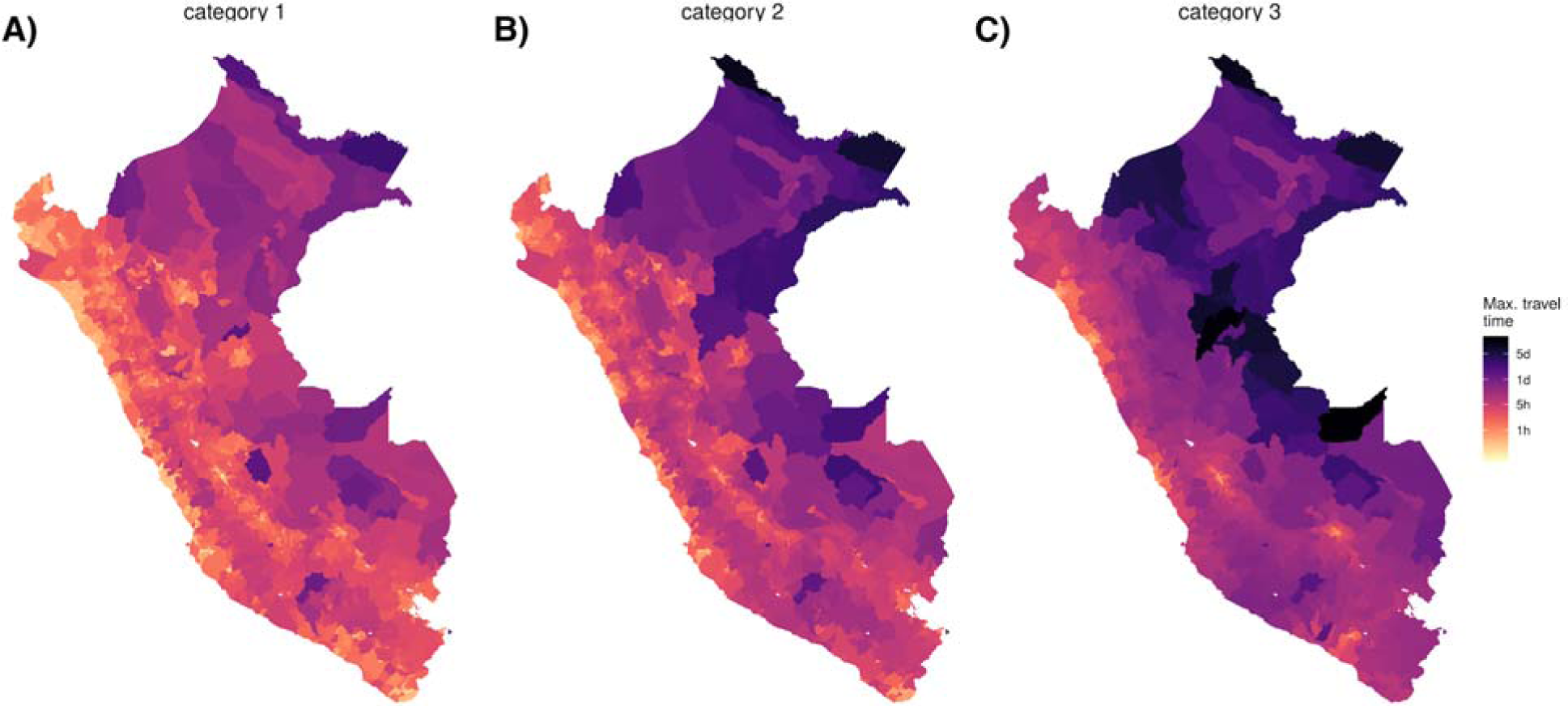
Country-wide map of travel time to health facilities for 2018. District-level average travel time to each category of healthcare facilities. A) Primary healthcare. B) Secondary healthcare. C) Tertiary healthcare. Color scale in logarithmic scale

### Urban/rural and ecological settings

High heterogeneity was observed in contrasting land coverage areas. The median travel time was 5.3 fold higher in rural (85 minutes; IQR=11–7,819) than in urban settings (16 minutes; IQR = 11–835) to a primary healthcare facility; 3.2 fold higher in rural (226 minutes; IQR = 11–12,429) than in urban settings (70 minutes; IQR = 11–3,386) to a secondary healthcare facility; and 2.4 fold higher in rural (568 minutes; IQR = 11–35,753) than in urban settings (235 minutes; IQR = 11–10,048) to a tertiary healthcare facility. A larger variation in travel time to primary healthcare was observed in rural compared to urban areas, and conversely, a larger variation in travel time to tertiary healthcare was observed in urban compared to rural areas (**Figure 3**). The district-level stratified averages in **Figure 2** show that there were also strong heterogeneities within major ecological regions. The north-east part of the Amazon Region, which borders with Colombia and Brazil, presented the largest country-wide travel times to the most proximate health facilities. The largest travel times to the most proximate health facilities within the Highlands Region was observed in the southern areas of the Andes, and in the coast was observed in the southern coast. Contrasting distributions of travel time to the most proximate health facility was observed between the 16 eco-regions defined by the MEnv (**Figure 3**).

**Figure 3.**
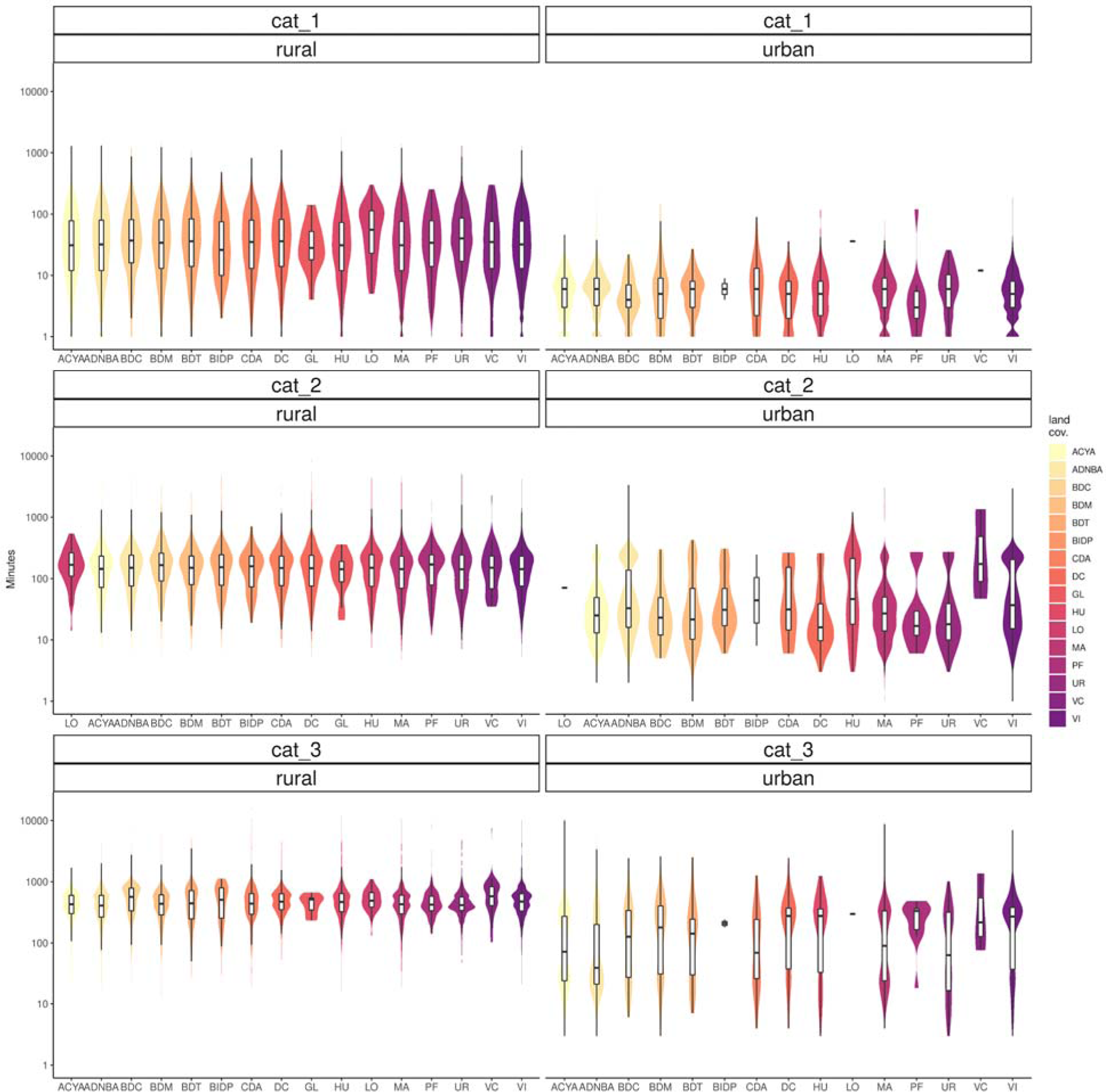
Distribution of travel time to most proximate health facility. Estimates across the 16 eco-regions defined by the Peruvian Ministry of environment and rural/urban settings for primary, secondary and tertiary healthcare. Y-axis in logarithmic scale.

### Travel time to health facilities relative to UBN

When the travel time to most proximate health facilities was distributed relative to the proportion of the population with unsatisfied basic needs at department level (administrative level 1), a positive trend was observed (**Figure 4**). The slope of this relation was increased in geographical accessibility to tertiary health facilities in comparison to primary or secondary health facilities.

**Figure 4.**
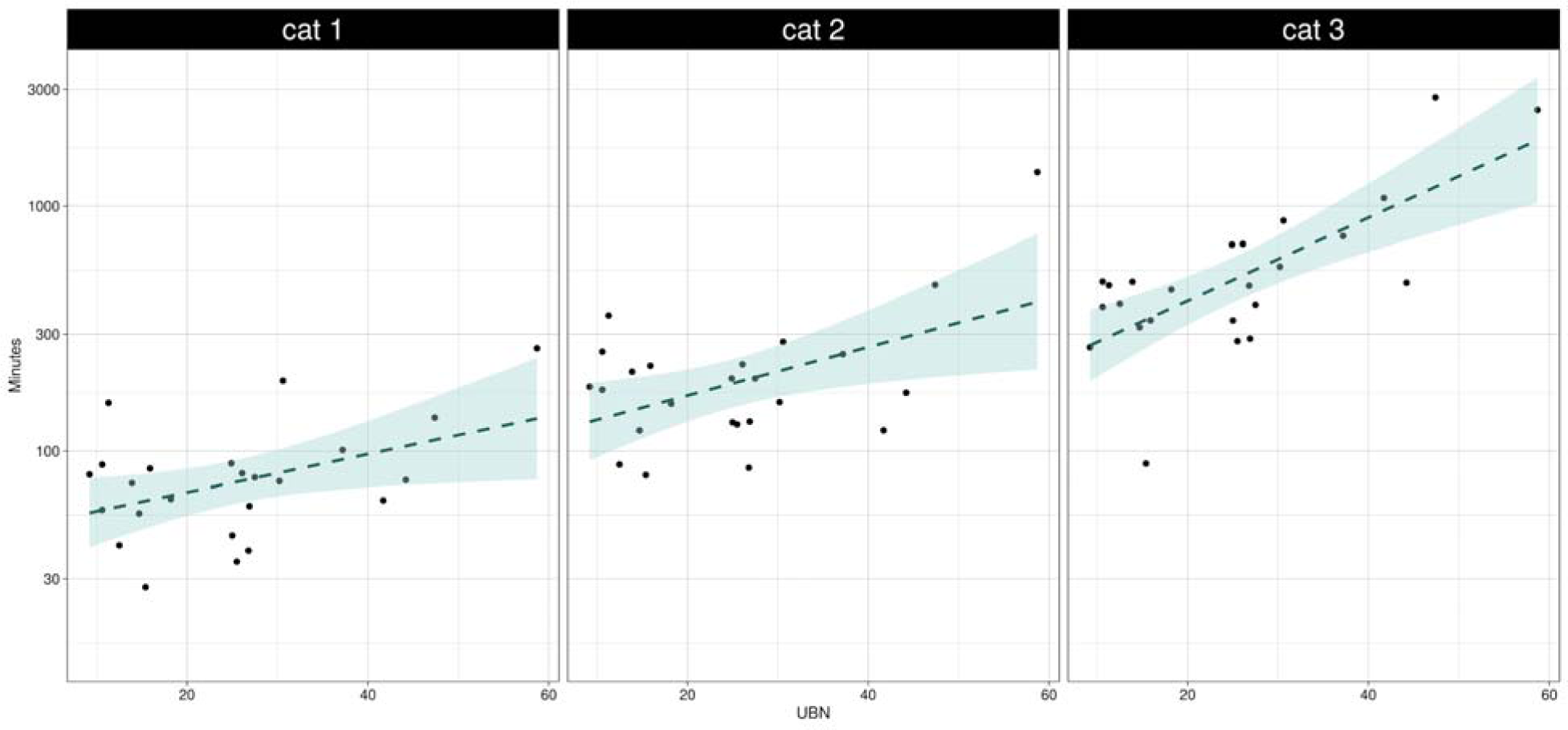
Median travel time to each health facility category relative to the proportion of population with unsatisfied basic needs per department. Y-axis in logarithmic scale

## 4. DISCUSSION

The present study explored the use of novel cloud-based geospatial modeling approach fitted with detailed local geospatial data to accurately estimate the travel time to the most proximate health facility across a highly diverse geographical and ecological settings as observed in Peru. This study showed the first quantification of heterogeneities in travel time to the most proximate health facility as a surrogate of geographical accessibility in the Latin American region. Most of the differences in travel time arose from heterogeneous land coverage profiles and the contrast between urban and rural areas. This is particularly important due to the fact that in Peru and in most LMIC, the most detailed data is available at a coarse administrative level that deter the resource planning and healthcare provision in these countries. Another direct implication of the utility of this approach is providing yet another angle of disadvantages amongst the most underserved, now in terms of access to healthcare as measured by distance and time, one of multiple aspects of high-quality healthcare.

In settings with a scattered distribution of villages, timely access to health facilities is a cornerstone to improve the health status of impoverished populations and a first step to provide high quality care (Kruk et al., 2018; Kruk, Pate, & Mullan, 2017). Although the use of big data and high-detail datasets paves the way for a comprehensive quantification of geographical accessibility in terms of distance and travel time, these technologies were not previously applied to estimate geographical accessibility to health facilities until recently (Tatem, 2018). Using this analytical approach, this study demonstrated that the population in the Jungle area have less accessibility since healthcare services are reachable at longer trajectories and travel time, understood as less geographical accessibility. The dramatic heterogeneity in travel time to the most proximate health facility observed in this study corresponds to the contrasting landscape composition in the coast, highlands, and jungle regions. A dense road network was observed in the Coast, facilitating access to multiple services including healthcare as reported in other studies in India and Africa (Kanuganti et al., 2015; Strano, Viana, Sorichetta, & Tatem, 2018).

Conversely, sparse road coverage was observed in the Highlands and only the two major cities in the Jungle region had roads.

Consistently with previous studies (Bashshur et al., 1971; Comber et al., 2011), this study determined the heterogeneity in travel time to the most proximate health facility across areas with contrasting land coverage types. Despite that this fact is widely accepted, few attempts have been made to quantify these heterogeneities. In addition, asymmetries were identified when the travel time to the most proximate health facility was compared along socio-economic profiles based on the unsatisfied basic needs index proposed by the United Nations Development Programme (UNDP). Uneven trends of greater travel time to health facilities (lower geographical accessibility) were observed among villages with higher rates of unmet basic needs. These results are consistent with previous reports of negative trends in geographical access to healthcare facilities in low-income populations (Kiwanuka et al., 2008; Meyer, Luong, Mamerow, & Ward, 2013; Peters et al., 2008; Tanser, Gijsbertsen, & Herbst, 2006).

It is important to highlight that the analysis conducted in this study did not take into account variability due to climatic factors that may prevent displacement to health facilities (i.e. floods or landslides). However, Highlands and Jungle areas are more prone to this kind of natural disaster, leading to a conservative estimation of travel time in these areas. Traffic, which may greatly influence the estimates in the large cities, was not considered in the analysis and potentially cause an underestimation of the travel time to health facilities. In addition, seasonal variability may greatly affect some displacement routes such as rivers; however, only navigable rivers were considered in this approach and the availability to displace through this rivers are less affected by seasonality. Another important consideration about the least-cost-path algorithm used in this analysis is that we infer the lowest travel time boundary to reach a health facility. This consideration relies on the assumption that the villagers opt for this route despite the cost and danger of the route in addition to its availability, as explained above.

In addition, the data reported here was generated at a meso-scale, with a spatial resolution of 30 meters. At this scale and resolution, some important details could be lost and affect the travel time estimations. For instance, in some settings the travel time might be increased due to meandering rivers or roads that follow the morphology of the terrain. The model assumes that transit flows in a direct manner, meaning that zigzagging routes may cause our approach underestimate the real travel time to reach a health facility. Despite these possible shortcomings, the proposed approach provided conservative yet useful estimates of travel times to health facilities that are important for planning of prevention and control strategies for multiple health-related events. This approach demonstrates that curation and alignment of geospatial data from multiple governmental institutions are important for national decision-making. In addition, the use of mapping and modeling techniques, and ‘big data’ were recognized as critical for better planning (Buckee et al., 2018; Hay, George, Moyes, & Brownstein, 2013; Tatem, 2018); however, a remaining challenge is the implementation of these approaches into routine disease prevention and control programs (Buckee et al., 2018; Hay et al., 2013).

This study acknowledges the relevance of other components of health access that may play an important role in the underlying phenomena. The sole presence of clinic infrastructure does not assure a proper healthcare delivery. Supply chain, human resources, financial accessibility, acceptability of services, and availability of treatment are some remaining barriers once geographical accessibility is overcome (Agbenyo et al., 2017; Al-Taiar et al., 2010; Johar, Soewondo, Pujisubekti, Satrio, & Adji, 2018). Further studies were suggested to get a comprehensive understanding of the accessibility to healthcare in Peru and other LMIC.

## 5. CONCLUSION

This study used a new methodology to estimate the travel time to most proximate health facilities as a first step to understanding and characterizing the geographical accessibility profiles in Peru. Contrasting patterns were observed across heterogeneous land coverage areas and urban and rural settings. These findings are important as first steps for tailoring strategies to deliver appropriate, affordable and quality healthcare to impoverished populations.

## Data Availability

Raw datasets and codes are available at google earth engine repository, details in the Supplementary Information section.

https://code.earthengine.google.com/1bf3524cfebcb89fa540e4c70b699c24

https://edgarmanrique30.users.earthengine.app/view/country-wide-map-of-travel-time-to-health-facilities-for-2018

## Acknowledgements

We thank the various ministries of Peru for making such useful data freely-available to researchers. Gabriel Carrasco-Escobar was supported by NIH/Fogarty International Center Global Infectious Diseases Training Program (D43 TW007120). J. Jaime Miranda acknowledges having received support from the Alliance for Health Policy and Systems Research (HQHSR1206660), the Bernard Lown Scholars in Cardiovascular Health Program at Harvard T.H. Chan School of Public Health (BLSCHP-1902), Bloomberg Philanthropies, FONDECYT via CIENCIACTIVA/CONCYTEC, British Council, British Embassy and the Newton-Paulet Fund (223-2018, 224-2018), DFID/MRC/Wellcome Global Health Trials (MR/M007405/1), Fogarty International Center (R21TW009982, D71TW010877), Grand Challenges Canada (0335-04), International Development Research Center Canada (IDRC 106887, 108167), Inter-American Institute for Global Change Research (IAI CRN3036), Medical Research Council (MR/P008984/1, MR/P024408/1, MR/P02386X/1), National Cancer Institute (1P20CA217231), National Heart, Lung and Blood Institute (HHSN268200900033C, 5U01HL114180, 1UM1HL134590), National Institute of Mental Health (1U19MH098780), Swiss National Science Foundation (40P740-160366), Wellcome (074833/Z/04/Z, 093541/Z/10/Z, 107435/Z/15/Z, 103994/Z/14/Z, 205177/Z/16/Z, 214185/Z/18/Z) and the World Diabetes Foundation (WDF15-1224). The funders had no role in study design, data collection and analysis, decision to publish, or preparation of the manuscript.

## Author’s contributions

Conceived and designed the study: G.C.E. and J.J.M. Data collection: G.C.E., K.T.L., and E.M. Analyzed the data: G.C.E., K.T.L., and E.M. Wrote the manuscript: G.C.E. and J.J.M. All authors reviewed and approved the final manuscript.

## Data Availability

Raw datasets and codes are available at google earth engine repository, details in the Supplementary information section.

